# Clinical evaluation of BD Veritor™ SARS-CoV-2 and Flu A+B Assay for point-of-care (POC) System

**DOI:** 10.1101/2021.05.04.21256323

**Authors:** Katherine Christensen, Huimiao Ren, Shirley Chen, Charles K. Cooper, Stephen Young

## Abstract

Differential diagnosis of COVID-19 and/or influenza (flu) at point of care is critical for efficient patient management and treatment for either of these diseases. Clinical performance of the BD Veritor™ System for Rapid Detection of SARS-CoV-2 & FluA+B (“Veritor SARS-CoV-2/Flu”) triplex assay was characterized. The performance for SARS-CoV-2 detection was determined using two hundred and ninety-eight (298) specimens from patients reporting COVID-19 symptoms within 7 days from symptom onset (DSO) in comparison with Lyra® SARS-CoV-2 RT-PCR Assay (“Lyra SARS-CoV-2”). The Veritor SARS-CoV-2/Flu Assay met the FDA EUA acceptance criterion with 95% overall agreement for SARS-CoV-2 test when compared to Lyra SARS-CoV-2. The performance for Flu A and Flu B detection was determined using 75 influenza-positive and 40 influenza-negative retrospective specimens in comparison with the previously FDA cleared BD Veritor™ System for Rapid Detection of Flu A+B (“Veritor Flu”). The Veritor SARS-CoV-2/Flu also demonstrated 100% agreement with the Veritor Flu.

## INTRODUCTION

Coronavirus disease 2019 (COVID-19) and influenza (flu) are two potentially severe respiratory illnesses that cause morbidity and mortality worldwide. COVID-19 is the result of infection by SARS-CoV-2, which emerged at the end of 2019.[1] Since then, over 147 million COVID-19 cases and 3.11 million COVID-19-related deaths have been reported worldwide.[2] The highly contagious nature of SARS-CoV-2, and the lack of any natural immunity in the world’s population, coupled with the fact that there is no efficient treatment for COVID-19, resulted in a global pandemic and public health crisis starting in 2020 and continuing in 2021. In the US, more than 32 million COVID-19 cases and over 572,000 COVID-19 deaths have been recorded through late April 2021.[2] Influenza is caused by influenza viruses (e.g. Flu A and Flu B), which occur seasonally.[3] With the exception of the 2020-2021 respiratory virus season, there are an estimated 1 billion cases of influenza-like illness identified each year globally. Within those cases, 3-5 million are severe, and 29,000-655,000 eventually lead to flu-related deaths.[4, 5] In the US, the 2019-2020 flu season resulted in over 38 million cases involving symptomatic illness and approximately 22,000 deaths.[6]

Patients with COVID-19 often exhibit respiratory symptoms similar to flu, including fever, cough, fatigue, headache, and muscle aches.[7] Some COVID-19 cases exhibit additional symptoms, including loss-of-smell/taste and shortness of breath; progression to severe disease can result in the loss of cardiopulmonary function and death. Although several of the clinical symptoms for flu overlap with symptoms for COVID-19, the therapeutic approaches for each illness are significantly different. While anti-viral drugs, such as Tamiflu® or Xofluza®, are often given to influenza patients, remdesivir and corticosteroids are the primary medications to date that have been utilized to treat COVID-19.[8] The early and rapid differential detection for SARS-CoV-2 versus influenza viruses is an essential requirement in determining the proper treatment for patients with the potential for infection by either of these viruses.[8] Accurate diagnosis of SARS-CoV-2 and/or influenza should also reduce the unnecessary burden placed on the healthcare system, especially during the respiratory virus seasons (e.g. December to February in the US).[8]

Molecular testing, utilizing real-time polymerase chain reaction (RT-PCR) assay technology, is the standard of care for the detection of most respiratory viral infections.[9, 10] However, this technique can be relatively labor and time-consuming, and laboratories generally need to have the necessary infrastructure and training to perform the methodology.[9] In addition, the turn-around-time for molecular testing conducted in a core laboratory requires additional time for transporting specimens. Rapid point-of-care (POC) molecular testing has reduced the assay time to between 15-30 minutes, but the cost of these assays can be prohibitive.[9] For many decentralized health care settings, a rapid testing platform that supports non-invasive specimen collection, is easy to use, and at a lower cost, is necessary. Therefore, several rapid antigen tests were developed to provide a sensitive but less expensive alternative point-of-care assay.[11, 12]

The BD Veritor™ System for Rapid Detection of Flu A+B is a US FDA cleared, and the BD Veritor™ System for Rapid Detection of SARS-CoV-2 is a US FDA EUA authorized, antigen-based testing system for use in POC settings.[13, 14] The clinical performance of both tests has been demonstrated by comparing with reference PCR-based assays.[15, 16] However, a triplex testing platform, which allows for the simultaneous detection of SARS-CoV-2, influenza A and influenza B viruses from one specimen, should help reduce the workflow burden while providing a differential diagnosis between COVID-19 and influenza.[8] The objective of this study was to demonstrate the clinical efficacy of a new triplex test, the BD Veritor SARS-CoV-2 & Flu A+B assay, to detect all three viral targets.

## METHODS AND MATERIALS

### Specimens and assays

This study was conducted as part of a US Food and Drug Administration-Emergency Use Authorization (EUA) submission. Clinical performance data from the BD Veritor™ System for Rapid Detection of SARS-CoV-2 & Flu A+B (“Veritor SARS-CoV-2/Flu”; Becton, Dickinson and Company; BD Life Sciences – Integrated Diagnostics Solutions, Sparks, MD, USA) was compared to both the Lyra® SARS-CoV-2 RT-PCR Assay (“Lyra SARS-CoV-2”; QUIDEL, San Diego, CA, USA) and the BD Veritor™ System for Rapid Detection of Flu A+B (“Veritor Flu”; Becton, Dickinson and Company; BD Life Sciences – Integrated Diagnostics Solutions, Sparks, MD, USA). The BD MAX SARS-CoV-2 RT-PCR Assay (“MAX SARS-CoV-2”; Becton, Dickinson and Company; BD Life Sciences—Integrated Diagnostics Solutions, Sparks, MD, USA) was used to resolve specimens with discrepant results between Veritor SARS-CoV-2/Flu and Lyra SARS-CoV-2 tests. The Lyra testing was performed according to the manufacturer’s instructions for use at TriCore Reference Laboratories, while the Veritor testing was performed internally at Becton, Dickinson and Company (BD Life Sciences – Integrated Diagnostics Solutions, San Diego, CA and Sparks, MD). The study protocol was approved by the Advarra Institutional Review Board (IRB).

Subjects ≥18 years of age symptomatic for COVID-19 were enrolled within seven DSO at six different sites across the US (Table S1) for SARS-CoV-2 testing. Nasal swabs were collected in duplicate. Two hundred and ninety-eight (298) specimens were collected between October 16 and October 30, 2020. Twenty specimens encountered technical errors and were not used in the analysis. The study used data from two hundred and seventy-eight (278) subjects for the analysis.

All study operators performing the Veritor SARS-CoV-2/Flu assay were blinded to reference method results.

A separate set of retrospective specimens were utilized to evaluate the clinical performance of Veritor SARS-CoV-2/Flu for Influenza testing. Residual de-identified nasopharyngeal (NP) swabs collected in universal viral transport (UVT) media were obtained from qualified specimen vendors. These clinical remnants included 75 influenza-positive specimens, 40 influenza A positive, 35 influenza B positive, and 40 influenza-negative specimens from subjects ranging from ≤5 to ≥60 years of age. The specimens were tested in a randomized and blinded fashion. All specimens used in this research were residual de-identified samples available after all standard of care testing was completed.

### Data analysis

The primary outcome measures for this study were positive and negative percent agreement, PPA and NPA, respectively. Point estimates with 95% confidence interval [95% CI] were calculated using the Wilson score method for the Veritor SARS-CoV-2/Flu assay, when compared to each reference method. The US FDA-EUA authorization acceptance criterion for test sensitivity for SARS-CoV-2 detection is a point estimate ≥ 80% (PPA) when compared to RT-PCR approach.[17] The test sensitivity for Flu A and Flu B detection was determined to be in agreement with the US FDA cleared BD Veritor™ System for Rapid Detection of Flu A+B, which met the US FDA-EUA authorization acceptance criterion of a point estimate ≥95% (PPA) with a lower bound of the 95% CI of 85% when compared to PCR assay. The Cohen’s kappa coefficient was applied to gauge agreement between reference and index tests to classify results into mutually exclusive categories. K=(P_o_ ^-P^e)/1-P_e_ (<0, 0, and >0 indicating agreements worse than, no better or worse than, and better than that expected by chance). The data presented in this report met the criteria as defined by the FDA guidance for Veritor SARS-CoV-2/Flu against the reference assays. This article was prepared according to STARD guidelines for diagnostic accuracy studies reporting.[18] The data will be available upon request.

## RESULTS

This study enrolled 298 specimens from subjects with COVID-19 within 7 days of symptom onset. Twenty (20) were considered unevaluable specimens and excluded. The remaining 278 specimens were tested with the reference method for SARS-CoV-2, the Quidel Lyra SARS-CoV-2 assay. The reference method testing resulted in 60 positive and 218 negative SARS-CoV-2 specimens. The collection procedure at site D deviated from the original protocol and the integrity of that site’s specimens may have been compromised. Although a statistically significant difference between D site and the five other sites (p=0.059) was not observed when determining data poolability, results that both include and exclude data obtained by D site are reported. For all sites, Veritor SARS-CoV-2/Flu had PPA and NPA values of 86.7% [95%CI: 75.8, 93.1] and 99.5% [95%CI: 97.4, 99.9], respectively, for the detection of SARS-CoV-2 compared to the reference (Table 1). Excluding the D site, the Veritor SARS-CoV-2/Flu had PPA and NPA values of 91.5% [95%CI: 80.1, 96.6] and 99.5% [95%CI: 97.0, 99.9], respectively, for the detection of SARS-CoV-2. The 22-49 years-of-age group had the highest percentage positive ratio within all positive cases compared to other age groups by both reference and Veritor SARS-CoV-2/Flu tests (Table 2).

**Table 1.** Performance of the Veritor SARS-CoV-2/Flu assay for detection of SARS-CoV-2 compared to reference with and without D site.

| **Collection Site** |  | **SARS-CoV2<sup>a</sup>** |
| --- | --- | --- |
| **D Site** | **PPA** | 69.2% [42.4, 87.3] |
|  | **NPA** | 100.0% [89.8, 100] |
|  | **Veritor (+) / Ref (+)** | 9 |
|  | **Veritor (+) / Ref (-)** | 0 |
|  | **Veritor (-) / Ref (+)** | 4 |
|  | **Veritor (-) / Ref (-)** | 34 |
|  | **kappa** | 0.765 |
| **Other Sites** | **PPA** | 91.5% [80.1, 96.6] |
|  | **NPA** | 99.5% [97.0, 99.9] |
|  | **Veritor (+) / Ref (+)** | 43 |
|  | **Veritor (+) / Ref (-)** | 1 |
|  | **Veritor (-) / Ref (+)** | 4 |
|  | **Veritor (-) / Ref (-)** | 183 |
|  | **kappa** | 0.9316 |
| **All Sites** | **PPA** | 86.7% [75.8, 93.1] |
|  | **NPA** | 99.5% [97.4, 99.9] |
|  | **Veritor (+) / Ref (+)** | 52 |
|  | **Veritor (+) / Ref (-)** | 1<sup>b</sup> |
|  | **Veritor (-) / Ref (+)** | 8 |
|  | **Veritor (-) / Ref (-)** | 217 |
|  | **kappa** | 0.9001 |

**Abbreviations:** PPA, positive percent agreement; NPA, negative percent agreement
<sup>a</sup>Reference method was the Lyra SARS-CoV-2 RT-PCR assay.
<sup>b</sup>Analysis of the Veritor Analyzer raw data demonstrated this specimen's results to be very close to the assay cut-off.

**Table 2.** SARS-CoV-2 positivity distribution by reference method or Veritor SARS-CoV-2/Flu across age groups.

| **Age group** |  |  |
| --- | --- | --- |
| **Without D Site** | **Reference**<br>**n (%)** | **Veritor SARS-CoV-2/Flu**<br>**n (%)** |
| **18-21 years** 3.5% (n=8) | 5 (10.6%) | 4 (9.1%) |
| **22-49 years** 55.0% (n=127) | 26 (55.3%) | 23 (52.3%) |
| **50-59 years** 22.9% (n=53) | 6 (12.8%) | 7 (15.9%) |
| **60-69 years** 13.9% (n=32) | 6 (12.8%) | 6 (13.6%) |
| **70-79 years** 3.5% (n=8) | 3 (6.4%) | 3 (6.8%) |
| **>=80 years** 1.3% (n=3) | 1 (2.1%) | 1 (2.3%) |
| **Overall (N=231)** | **47** | **44** |
| **All Sites** |  |  |
| **18-21 years** 3.6% (n=10) | 5 (8.3%) | 4 (7.5%) |
| **22-49 years** 52.9% (n=147) | 31 (51.7%) | 25 (47.2%) |
| **50-59 years** 22.7% (n=63) | 10 (16.7%) | 11 (20.8%) |
| **60-69 years** 15.1% (n=42) | 7 (11.7%) | 7 (13.2%) |
| **70-79 years** 4.7% (n=13) | 6 (10.0%) | 5 (9.4%) |
| **>=80 years** 1.1% (n=3) | 1 (1.7%) | 1 (1.9%) |
| **Overall (N=278)** | **60** | **53** |

Discordant results were observed from nine out of the 278 total specimens (Table S2). Eight specimens positive by the Lyra SARS-CoV-2 assay were negative by the Veritor SARS-CoV-2/Flu assay. Two of the eight specimens were associated with Ct values of <30; the other six had Ct values ≥30 (Table S3). The BD MAX SARS-CoV-2 assay was used to resolve discordant results. Seven of the eight discordant specimens were positive by the BD MAX SARS-CoV-2 assay. The other one was negative by the BD MAX SARS-CoV-2 assay. One specimen positive by the Veritor SARS-CoV-2/Flu assay was negative by Lyra.

The clinical study was conducted in the early part of the 2020-2021 flu season, therefore, the concurrence of SARS-CoV-2 and influenza virus in this study was non-existent. Only one Flu A positive and two Flu B positives were reported by Veritor. One Flu B positive reported by Veritor was also shown as SARS-CoV-2 positive by both Veritor and Lyra reference results. These three specimens were tested on MAX and resulted as negative suggesting they were false positives for Flu A and Flu B.

Given the lack of prospective Flu A & B samples, the sensitivity of the Flu A and B detection, 75 retrospective residual de-identified positive influenza specimens (40 influenza A positive, 35 influenza B positive) and 40 negative influenza A/B remnant specimens were used to determine the clinical performance of Veritor SARS-CoV-2/Flu assay. The Veritor™ SARS-CoV-2/Flu results were further compared to results from the comparator method, the FDA cleared BD Veritor Flu A/B test. The results from this testing were used to determine PPA and NPA values (Table 3). For Flu A detection, Veritor SARS-CoV-2/Flu had PPA and NPA values of 100% [95%CI: 91.2, 100] and 100% [95%CI: 95.2, 100], respectively; for Flu B, Veritor SARS-CoV-2/Flu had PPA and NPA values of 100% [95%CI: 90.0, 100] and 100% [95%CI: 95.5, 100], respectively (Table 4). Age stratifying the positive samples resulted in the ≥60 years old group having the highest Flu A positivity ratio than other age groups (Table 4). However, most Flu B positive samples fell in the age group ranging from 6 to 59 years old.

**Table 3.** Performance of the Veritor SARS-CoV-2/Flu assay for detection of Flu A and Flu B compared to reference.

|  | **Flu A<sup>a</sup>** | **Flu B<sup>a</sup>** |
| --- | --- | --- |
| **PPA** | 100 [91.2, 100] | 100 [90.0, 100] |
| **NPA** | 100 [95.2, 100] | 100 [95.5, 100] |
| **Veritor (+) / Ref (+)** | 40 | 35 |
| **Veritor (+) / Ref (-)** | 0 | 0 |
| **Veritor (-) / Ref (+)** | 0 | 0 |
| **Veritor (-) / Ref (-)** | 75 | 80 |
| **kappa** | 1 | 1 |

**Abbreviations:** PPA, positive percent agreement; NPA, negative percent agreement
<sup>a</sup>Reference method was the BD Veritor System Flu A+B assay

**Table 4.** Influenza positivity distribution by reference method or Veritor SARS-CoV-2/Flu across age groups.

| Age group | Reference |  | Veritor SARS-CoV-2/Flu |  |
| --- | --- | --- | --- | --- |
|  | Influenza A<br>n (%) | Influenza B<br>n (%) | Influenza A<br>n (%) | Influenza B<br>n (%) |
| **≤5 years** 13.0% (n=15) | 3 (7.5%) | 8 (22.9%) | 3 (7.5%) | 8 (22.9%) |
| **6-21 years** 20.0% (n=23) | 6 (15%) | 13 (37.1%) | 6 (15%) | 13 (37.1%) |
| **22-59 years** 33.9% (n=39) | 11 (27.5%) | 13 (37.1%) | 11 (27.5%) | 13 (37.1%) |
| **≥60 years** 33.0% (n=38) | 20 (50.0%) | 1 (2.9%) | 20 (50.0%) | 1 (2.9%) |
| **Overall** (N=115) | 40 | 35 | 40 | 35 |

## DISCUSSION

The results presented here show PPA values for the Veritor SARS-CoV-2/Flu assay compared to a RT-PCR assay met FDA-EUA acceptance criteria for detection of SARS-CoV-2 (86.7%; [95%CI: 75.8, 93.1]). Similarly, the Veritor SARS-CoV-2/Flu assay demonstrated an NPA value of 99.5% for detection of SARS-CoV-2 compared to an RT-PCR reference method. Although a marginal statistically significant difference between D site and the five other sites (p=0.059) was shown, the PPA both including (86.7%) and excluding (91.5%) D site met the FDA-EUA acceptance criteria for detecting SARS-CoV-2. Additionally, the Flu detection portion of the Veritor SARS-CoV-2/Flu assay demonstrated 100% agreement with the 510k cleared BD Veritor System Flu A+B assay. For Flu A detection, the lower bound of the 95% CI was 91.2%, and for Flu B detection, the lower bound was 90.0%.

Veritor SARS-CoV2/Flu assay showed a reduced capacity to detect SARS-CoV-2 when the corresponding reference test result had Ct values ≥30 during discordant testing. This is a common observation for antigen tests since most assays rely on releasing the protein target that can flow by capillary action to initiate the antibody-antigen complex and the detection reaction. Therefore, viable viral particles are required for antigen detection.[19] In contrast, PCR-based assays detect viral nucleic acid, reflecting viral shedding but not active infection. Viral load and analytical sensitivity of the reference RT-PCR assay heavily influence the sensitivity of the antigen test.[19, 20] Thus, RT-PCR-based assays may seem more sensitive, but they do not necessarily reflect infectivity of COVID-19; whereas, antigen testing is a more specific approach for SARS-CoV-2 screening (compared to RT-PCR) and aligns with infectiousness of the tested individual.[21]

The performance of Veritor SARS-CoV-2/Flu for Flu A and Flu B showed 100% PPA with the reference method, Veritor Flu, suggesting the same sensitivity for Flu detection. All 278 specimens tested for COVID-19 returned only one positive for Flu A and two positives for Flu B. These three specimens were further determined as false positives for Flu A and Flu B. This could be due to a higher infection rate for SARS-CoV-2 or to the low activity of flu during the time of specimen collection (October, 2020).[22] The 2020-2021 respiratory virus season will conclude with an extremely low prevalence of influenza-like illness. However, in a season when the incidence of both COVID-19 and flu cases is high, the differential diagnosis of each agent for the appropriate therapeutic approach could be less challenging by using the Veritor SARS-CoV-2/Flu assay. Especially, when the currently available medications for treating both illnesses differ, and the treatment indications for the diseases do not overlap. Safety concerns for treatment could arise if false positive or false negative results occur between the two infections.

Additionally, proper quarantine and contact tracing are essential steps for preventing the spread of the infectious disease. Although COVID-19 and influenza share a similar transmission mechanism and have overlapping clinical symptoms, the quarantine length and the therapeutic approach for each illness are not the same.[23] After symptoms onset, the recommended quarantine period is a minimum of 4-5 days for flu,[24] but a minimum of 10 days for COVID-19.[25] Therefore, the accurate detection of both SARS-CoV-2 and Flu A+B impacts not only the treatment plan but also the period of quarantine and resulting loss of work and school attendance. The Veritor SARS-CoV-2/Flu test could provide comparable clinical outcomes to the molecular testing approach by speeding up the diagnosis result to guide and initiate the correct and timely therapeutic approach.

Different technologies are currently available for detecting SARS-CoV-2 and Flu A/B viruses for the diagnosis of COVID-19 and flu, respectively.[12, 19] Although the RT-PCR-based approach currently represents the laboratory method of choice due to its relatively high analytic and clinical sensitivity, rapid tests carry several advantages, including faster turnaround time and more straightforward implementation in decentralized health care settings for POC purposes.[10, 12] Depending on the infrastructure and available resources in the health care facility, the BD Veritor SARS-CoV-2/Flu assay could aid the process of distinguishing the detection and diagnosis of COVID-19 and flu for proper patient triage, disease mitigation, and managing treatment.

### Limitations

The test for Flu A and Flu B was conducted by using materials obtained from pre-selected frozen remnants. Unbiased subjects with no confirmed diagnosis should be considered for testing.

### Conclusions

The Veritor SARS-CoV-2/Flu assay met US FDA-EUA acceptance criteria for SARS-CoV-2 detection. The test sensitivity of the Veritor SARS-CoV-2/Flu assay for Flu A and B detection was in agreement with the previously cleared Veritor System Flu A+B assay. Dual detection capability for the etiologic agents causing COVID-19 and influenza will allow efficient differentiation between the two illnesses and will inform physicians regarding diagnosis and, therefore, the proper treatment and disease management for patients exhibiting similar symptoms. Dual testing may be especially important for the duration of the COVID-19 pandemic as it overlaps with flu season and could have a major impact in decentralized health care settings.

## Data Availability

The data will be available upon request.

## ACKNOWLEDGEMENTS

We thank Karen Eckert and Yu-Chih Lin for their input on the content of this manuscript. We thank Yongqiang Zhang (Becton, Dickinson and Company, BD Life Sciences – Integrated Diagnostic Solutions) for statistical support. We are grateful to the study participants who allowed this work to be performed.

## AUTHOR CONTRIBUTIONS

**Katherine Christensen:** Conceptualization, Methodology, Investigation, Resources, Writing – Original Draft, Visualization, Project administration. **Huimiao Ren:** Methodology, Investigation, Formal analysis, Data Curation, Writing – Review & Editing. **Shirley Chen:** Methodology, Investigation, Formal analysis, Data Curation, Writing – Review & Editing. **Charles K. Cooper:** Conceptualization, Writing – Review & Editing, Supervision, Funding acquisition. **Stephen Young:** Conceptualization, Investigation, Resources, Writing – Review & Editing, Supervision, Funding acquisition.

---

**Table S1.** SARS-CoV-2 specimens collection sites.

| **Site** | **City** | **State** | **Enrollment** |
| --- | --- | --- | --- |
| A | Palm Springs | FL | 46 |
| B | Savannah | GA | 94 |
| C | Ft. Lauderdale | FL | 80 |
| D<sup>a</sup> | Downers Grove | IL | 64 |
| E | Chattanooga | TN | 11 |
| F | St. Louis | MO | 3 |
| **TOTAL** |  |  | **298** |

<sup>a</sup>The collection procedure was not consistent with the protocols utilized by other sites. The collection deviation was noted and therefore, the data was reported with and without this site.

**Table S2.** List of discordant specimens.

| **Specimen ID** | **Veritor** | **Lyra** | **MAX** | **MAX Ct** |
| --- | --- | --- | --- | --- |
| 1 | - | + | + | N1: 29.06, N2: 29.48 |
| 2 | - | + | + | N1: 26.38, N2: 29.48 |
| 3 | - | + | - | N/A |
| 4 | - | + | + | N1: 30.79, N2: 32.72 |
| 5 | - | + | + | N1: 30.43, N2: 32.38 |
| 6 | - | + | + | N1: 38.27, N2: 32.71 |
| 7 | - | + | + | N1: 31.82, N2: 33.95 |
| 8<sup>a</sup> | + | - | - | N/A |

<sup>a</sup>Analysis of the Veritor Analyzer raw data demonstrated this specimen's results to be very close to the assay cut-off.

**Table S3.** Comparison of Veritor SARS-CoV-2/Flu assay results with those from the Lyra SARS-CoV-2 assay, stratified by cycle threshold category, with and without D site.

| **Without D Site** |  | **Lyra SARS-CoV-2** |
| --- | --- | --- |
| **Veritor SARS-CoV-2** | **Positive (Ct ≤ 30)** | **Positive (Ct > 30)** |
| **Positive** | 42 | 1 |
| **Negative** | 0 | 4 |
| **Total** | 42 | 5 |
| **PPA (95% CI)** | 100% (91.6% - 100%) | 20.0% (3.6% - 62.4%) |

| **All Sites** |  | **Lyra SARS-CoV-2** |
| --- | --- | --- |
| **Veritor SARS-CoV-2** | **Positive (Ct ≤ 30)** | **Positive (Ct > 30)** |
| **Positive** | 50 | 2 |
| **Negative** | 2 | 6 |
| **Total** | 52 | 8 |
| **PPA (95% CI)** | 100% (92.6% - 100%) | 100% (74.1% - 100%) |

**Abbreviations:** Ct, PCR cycle threshold; PPA, positive percent agreement

## REFERENCES

1. Zhu, N., et al., A Novel Coronavirus from Patients with Pneumonia in China, 2019. N Engl J Med, 2020. 382(8): p. 727–733.

2. Johns Hopkins University and Medicine, Coronavirus Resource Center, Mortality Analyses. 2020. https://coronavirus.jhu.edu/data/mortality.

3. Centers for Disease Control and Prevention, Influenza (Flu). Accessed December 16, 2020. https://www.cdc.gov/flu/about/index.html.

4. Iuliano, A.D., et al., Estimates of global seasonal influenza-associated respiratory mortality: a modelling study. Lancet, 2018. 391(10127): p. 1285–1300.

5. World Health Organization, Global influenza strategy 2019-2030. Geneva: 2019. Licence: CC BY-NC-SA 3.0 IGO. Cataloguing-in-Publication (CIP) data. CIP data are available at http://apps.who.int/iris.

6. Centers for Disease Control and Prevention, Estimated Influenza Illness, Medical Visits, Hospitalizations, and Deaths in the United States-2019-2020 Influenza Season. https://www.cdc.gov/flu/about/burden/2019-2020.html. 2020.

7. Centers for Disease Control and Prevention, Similarites and differences between Flu and COVID-19. Accessed December 28, 2020. https://www.cdc.gov/flu/symptoms/flu-vscovid19.htm#:∼:text=Flu%20viruses%20can%20cause%20mild,signs%20and%20symptoms%20listed%20above.&text=COVID%2D19%20seems%20to%20cause,loss%20of%20taste%20or%20smell.

8. Rubin, R., What Happens When COVID-19 Collides With Flu Season? JAMA, 2020. 324(10): p. xs923–925.

9. Yarbrough, M.L., et al., Influence of Molecular Testing on Influenza Diagnosis. Clinical chemistry, 2018. 64(11): p. 1560–1566.

10. Cheng, M.P., et al., Diagnostic Testing for Severe Acute Respiratory Syndrome–Related Coronavirus 2. Annals of Internal Medicine, 2020. 172(11): p. 726–734.

11. World Health Organization, Global Research Collaboration for Infectious Disease Preparedness. COVID 19: Public Health Emergency of International Concern (PHEIC). Global Research and Innovation Forum: Towards a Research Roadmap. 02/11/2020-02/12/2020. https://www.who.int/blueprint/priority-diseases/key-action/Global_Research_Forum_FINAL_VERSION_for_web_14_feb_2020.pdf?ua=1.

12. Vemula, S.V., et al., Current Approaches for Diagnosis of Influenza Virus Infections in Humans. Viruses, 2016. 8(4): p. 96–96.

13. BD Veritor™ System for Rapid Detection of Flu A+B [package insert, EUA]. Becton, Dickinson and Company, Sparks-Glencoe, MD; (2018).

14. BD Veritor™ System for Rapid Detection of SARS-CoV-2 [package insert, EUA]. Becton, Dickinson and Company, Sparks-Glencoe, MD; (2020).

15. Nam, M.H., et al., Clinical performance evaluation of the BD Veritor System Flu A+B assay. J Virol Methods, 2014. 204: p. 86–90.

16. Young, S., et al., Clinical Evaluation of BD Veritor SARS-CoV-2 Point-of-Care Test Performance Compared to PCR-Based Testing and versus the Sofia 2 SARS Antigen Point-of-Care Test. J Clin Microbiol, 2020. 59(1).

17. U.S. Food and Drug Administration, Coronavirus Disease 2019 (COVID-19) Emergency Use Authorization for Medical Devices--In Vitro Diagnostics EUAs.https://www.fda.gov/medical-devices/coronavirus-disease-2019-covid-19-emergency-use-authorizations-medical-devices/vitro-diagnostics-euas#individual-molecular.

18. Bossuyt, P.M., et al., STARD 2015: An Updated List of Essential Items for Reporting Diagnostic Accuracy Studies. Radiology, 2015. 277(3): p. 826–32.

19. La Marca, A., et al., Testing for SARS-CoV-2 (COVID-19): a systematic review and clinical guide to molecular and serological in-vitro diagnostic assays. Reprod Biomed Online, 2020. 41(3): p. 483–499.

20. Diao, B., et al., Diagnosis of Acute Respiratory Syndrome Coronavirus 2 Infection by Detection of Nucleocapsid Protein. medRxiv, 2020: p. 2020.03.07.20032524.

21. Pekosz, A., et al., Antigen-Based Testing but Not Real-Time Polymerase Chain Reaction Correlates With Severe Acute Respiratory Syndrome Coronavirus 2 Viral Culture. Clin Infect Dis, 2021.

22. Petersen, E., et al., Comparing SARS-CoV-2 with SARS-CoV and influenza pandemics. The Lancet Infectious Diseases, 2020.

23. Kaur, S.P. and V. Gupta, COVID-19 Vaccine: A comprehensive status report. Virus research, 2020. 288: p. 198114–198114.

24. Centers for Disease Control and Prevention, Stay at home when you are sick. Accessed December 28, 2020. https://www.cdc.gov/flu/business/stay-home-when-sick.htm#:∼:text=Individuals%20with%20suspected%20or%20confirmed,3%20days%20of%20their%20illness.

25. Centers for Disease Control and Prevention, Options to reduce quarantine for contacts of persons with SARS-CoV-2 infection using symptom monitoring and diagnostic testing. Accessed December 28, 2020. https://www.cdc.gov/coronavirus/2019-ncov/more/scientific-brief-options-to-reduce-quarantine.html.

